# Secondary Household Covid-19 Transmission Modelling of Students Returning Home from University

**DOI:** 10.1101/2020.11.11.20229559

**Authors:** Paul R. Harper, Joshua W. Moore, Thomas E. Woolley

## Abstract

We estimate the number of secondary Covid-19 infections caused by potentially infectious students returning from university to private homes with other occupants. Using a Monte-Carlo method and data derived from UK sources, we predict that an infectious student would, on average, infect 0.94 other household members. Or, as a rule of thumb, each infected student would generate (just less than) one secondary within-household infection. The total number of secondary cases for all returning students is dependent on the virus prevalence within the student population at the time of their departure from campus back home. Correspondingly, we provide results for prevalence ranging from 0.5% to 15%, which is based on observed minimum and maximum estimates from Cardiff University’s asymptomatic testing service. Although the proposed estimation method is general and robust, the results are sensitive to the input data. We therefore provide Matlab code and a helpful online app (http://bit.ly/Secondary_infections_app) that can be used to estimate numbers of secondary infections based on local parameter values.

## I. INTRODUCTION

SARS-CoV-2, otherwise known as Covid-19, or corona virus, has caused a global pandemic lasting throughout 2020 [1–3]. Governments tried to stem the infection spread by imposing national lockdowns, resulting in many services, such as education, rapidly transitioning to online spaces [4].

Concerns had been raised [5, 6] about potential high rates of transmissions between university students as they were encouraged to return to campus, especially those living together in large capacity, close confinement, housing, such as university halls of residence. Indeed, many universities initially reported localised surges in infections [7] confirming indications that infections are more likely to occur due to contact with peers rather than across age bands [8].

It could been suggested that since young people (the typical age of a student) generally recover well from covid-19 [9] then allowing them to pass the disease rapidly between themselves is actually a good way of producing herd immunity, at least for their age group [10]. However, questions regarding how long the immunity will last [11–14] coupled with suffering through the actual disease and the stress of missing coursework [15] leads this to be an unsympathetic plan of action.

Critically, even if herd immunity was possible in the university student body the students are going to return to their private domiciles at the end of term and pose a risk to everyone in their household. Although there has been a focus on modelling the future trajectories of an infection outbreak within a university [16], our work explores an area that has seemingly not been previously considered: namely, what is the impact if students return home during lockdown, or at the end of term? Even if students are able to recover well, or even be asymptomatic, they can still be carriers [8]. Thus, they can take the infection into a household that may contain other adults of different age groups (*e*.*g*. parents) that may not be expected to recover as well, particularly if they have an underlying long term illness. Further, this is a timely question to ask due to the winter holiday break being a major time for family gatherings resulting in a higher risk of extended contacts with cross-sectional age groups.

To estimate the number of secondary infections due to university student returning home we apply a Monte-Carlo approach to UK-based data. Namely, We combine the prevalence of infection at university (in this case Cardiff University), with distributions of household size and probability of secondary infections, to predict the mean expected number of secondary infections, as well as 95% confidence intervals.

Although our results are based on UK data we provide the reader with Matlab code (Appendix A) and an online app (http://bit.ly/Secondary_infections_app) that not only reproduces our results, but can also be adapted by the reader to include data which is more accurate and/or specific to their location and needs. The full Matlab and app codes are kept in an online repository (http://bit.ly/secondary_infections_repo) and will be updated as required.

The user’s local data and subsequent results can be used to develop and evidence a university’s strategy for dealing with an infected student body and their desire to return home for the holidays. To this end after providing our results and conclusions we offer a number of strategies for universities to consider, highlighting each scheme’s positive and negative points. Our hope is that our work will enable universities to take action as soon as possible, based on the best predictive estimates we can offer.

## II. METHODS AND DATA SOURCES

We define:

- *I* to be the prevalence of the virus in the student body. Specifically, this is the percentage probability that an individual student has Covid-19 and is infectious. In this study this is as a control parameter, which is informed by data from [17, 18]. See Section II B.
- *S* to be the probability of a secondary transmission to another member of the household. This is a continuous random variable, assumed to be normally distributed. The mean and standard deviation is informed by data from [19]. See Section II C.
- *H* to be the number of occupants in a household, other than the student themselves. This is a discrete random variable whose distribution is informed by [20]. See Section II D.
- *N* to be the total number of students returning back home from campus. Here, we take *N* = 1000, so the results can be stated “per thousand students”.

The total number of additional (secondary) household infections, *T*, is thus calculated by:

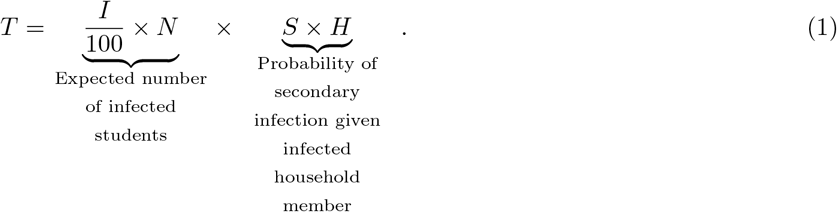

Although using the average values of *S* and *H* would provide a “rule of thumb” measure for *T*, we have decided to extend this investigation and use a Monte-Carlo approach to sample from the specified distributions of *S* and *H* [21]. Not only does this method allow us to capture the stochastic nature of virus transmission, but equally, it will provide us with error bounds on the number of predicted secondary infections.

### A. Monte-Carlo algorithm

The central idea of the algorithm rests on sampling *N* times from the household size and secondary infection probability distributions to generate approximations for *H*_*k*_ and *S*_*k*_ *k* = 1, …, *N*, respectively. We present a general method for sampling from these distributions, which can be coded up in a users’ language of choice. Alternatively, we provide a full working Matlab code in Appendix A, or for those who want a quick result without resorting to coding we offer an online app, which can be accessed at http://bit.ly/Secondary_infections_app.

The household size probabilities form a discrete distribution. Specifically, we define *p*_*i*_ to be the probability of there being 1 ≤ *i* ≤ *n* people in the household other than the student. For clarity, we explicitly state that *p*_*i*_ would be the proportion of households containing *i* + 1 people, *i*.*e*. the infected student and the *i* ‘other’ members of the household. We note that in our current case *n* = 5.

To sample from this distribution we generate a uniformly distributed random number, *r* and the susceptible household size, is defined to be *j* such that

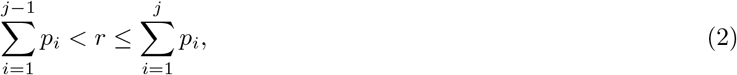

where we note that *p*_0_ = 0 because we are only considering the cases where students are returning to households containing potential susceptible people. This will lead to a small over estimate as we are ignoring any student who lives on their own outside of term time, however, we assume such cases are in a small minority. Finally, we denote *H*_*k*_ = *j* to be the *k*th family size.

From the data on secondary infections [19] we are supplied with the mean probability of secondary infection, *µ*_*i*_, and 95% confidence intervals for households of size *i* = 2, 3, …, 6. We assume that the probability is normally distributed and convert the 95% confidence interval back into a standard deviation, *σ*_*i*_. We then sample *N* times from these normal distributions, 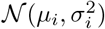, for each family size, *i*, capping the probability at zero and one. Finally, we denote *S*_*k*_(*i*) to be this sampled value, *i*.*e*. the *k*th probability of secondary infection in a household of size *i*.

Finally, we simulate *N* Bernoulli trials with probability of “success” given by *I/*100 and defining *I*_*k*_ = 1 if the simulation is “succesful” and *I*_*k*_ = 0 if not. Here, *I*_*k*_ is acting as an indicator variable that the *k*th household is home to an infected student.

The total number of secondary infected cases is then given by

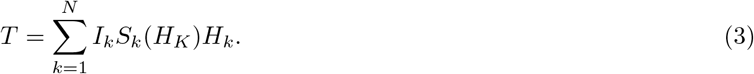

The above procedure can be repeated as many times as required to resolve the statistics to any required accuracy. Further, it can be observed that in the case where we collapse the distributions onto their mean values equation (3) simplifies to equation (1).

### B. Probability that an individual student has Covid-19 and is infectious when returning home, *I*

Obtaining an accurate value of *I* is problematic due to it evolving over time and varying across the UK. Specifically, *I* is influenced by many factors, such as local and national lockdown measures and adherence to social distancing. To include such variance we present the total number of new infections, *T*, for a range of different values of *I* that reflect a range of localised prevalences possible at the time of departure (when students return home).

Recent ONS data [17] suggests that for the age band 12-24, *I* = 1.5% *i*.*e*. 1.5% of those in that age category are infected. However, case data from Cardiff University’s asymptomatic testing service [18] indicated that the underlying rate may have been as high as *I* = 15% (w/c 10th October 2020) but at the time of writing the rate has dropped to well under *I* = 5%. To encompass this range we use *I* ∈ *{*0.5, 1.5, 5, 10, 15*}*.

### C. Secondary transmission probabilities, *S*

Several publications, for example [22–24] have reported estimates for *S*. We have used those in a more recent UK-wide study [19] given the UK context and that, reassuringly, the estimate is broadly in-line with other studies. Overall, the authors report the probability of a secondary infection *S* = 0.37 i.e. 37% (95% CI 31-43%). In this Monte-Carlo study we use the more detailed information from [19], where the authors are able to differentiate values of *S* by household size. The data set can be found in Table I.

**TABLE I:** Probability of secondary infections occurring in households of different sizes. Adapted from Lopez Bernal et al. [19].

| Household size | Probability of secondary infection<br>with 95% confidence interval |
| --- | --- |
| 2 | 0.49 (0.37 - 0.60) |
| 3 | 0.41 (0.29 - 0.52) |
| 4 | 0.32 (0.22 - 0.42) |
| $\geq 5$ | 0.25 (0.14 - 0.36) |

We note that in households of size 2, *S* = 0.49 (95% CI 0.37 – 0.61) i.e. a Covid-19 positive student has, on average, a 49% chance of making the other occupant infected. However, for houses with 5 or more occupants, this reduces to *S* = 0.25 (25%) for each occupant. It is unexpected that the probability should decrease with an increased household size. To account for this, we suggest that this may be an actual result that shows that larger households are more likely to be more cautious in their interactions. Alternatively, this may simply be a problem with the low number of recordings in the data. Namely, Covid-19 is, thankfully, rare and household sizes ≥ 5 are also rare (see Section II D), thus the *S* = 0.25 value is derived from only ∼ 30 reported cases of secondary infections occurring in such households.

### D. Household size, *H*

Data on the number and proportion of households in Wales containing at least one student (2019) has been provided by Economic and Labour Market Statistics, Welsh Government, and is summarised below. The average household size can be calculated to be 3.2 people, however, we use the full distribution provided by the data in the Monte-Carlo simulation, as described in Section II A. Specifically, each household size is linked to different values of the probability of secondary transmissions, *S*.

## III. CODES AND APP

Alongside the Matlab code in Appendix A that can be used to reproduce our results, as well as be adapted to a user’s specific data set, we have also developed an online app, which can be found at http://bit.ly/Secondary_infections_app, for ease of use. Full Matlab and app codes will be kept in an online repository, http://bit.ly/secondary_infections_repo.

Figure 1 illustrates the user interface. The top contains a table of values which can be edited by the user. The values in the table must all be positive and less than one. Further, the sum of the probabilities along the top ‘household’ row, must sum to less than one. Next the sliders on the left are set to the chosen number of students to be simulated, *N*, and the chosen prevalence rate, *I*.

**FIG. 1:**
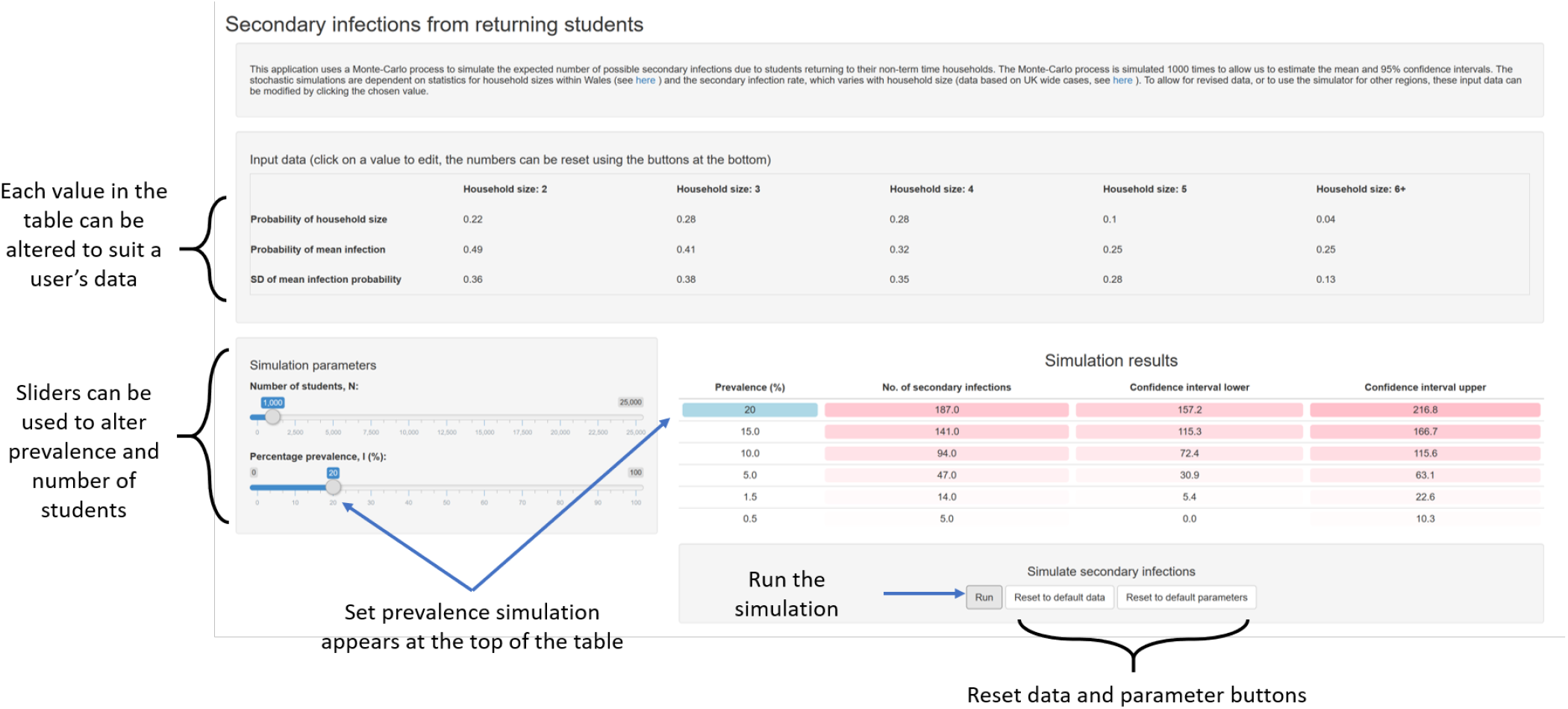
Online app interface. The app can be found at http://bit.ly/Secondary_infections_app.

Once these values are set, the ‘Run’ button below the table on the right should be clicked and after a few seconds of simulation the results table will be filled with predicted results. Namely, the top row corresponds to the user’s chosen prevalence rate and presents the mean number of secondary infections, as well as the 95% confidence intervals for the prevalence rates. The rows below this present the prevalence rates for *I* ∈ {0.5, 1.5, 5.0, 10.0, 15.0}, which are used to compare with the results presented in this paper.

## IV. RESULTS

Figure 2 presents histograms of 10^4^ Monte-Carlo stochastic trials calculating *T* from equation (3), for varying prevalence rates. Due to the central limit theorem, the assumption that the final distribution is Gaussian is more applicable to the simulations with higher prevalence rates, *i*.*e*. when more people are infected. However, even in the low prevalence case of *I* = 0.5% the Gaussian fit is easily within the error tolerances of the data we have.

**FIG. 2:**
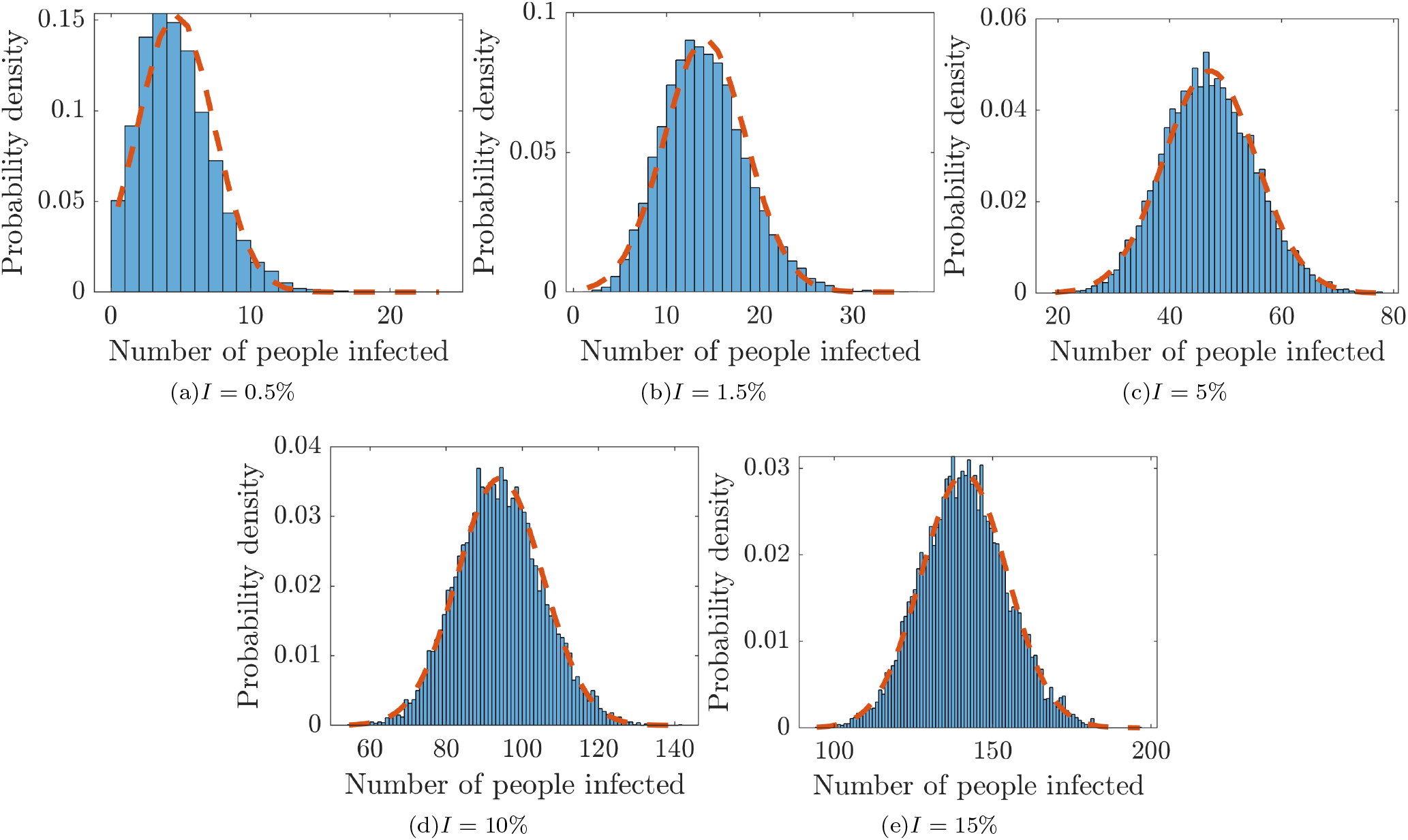
Histogram and data fit representing the variability over 10^4^ Monte-Carlo simulations. The blue histogram is the simulated data, the red dashed line is the fit of a normal distribution. Note that we are plotting the probability density, but since each column is one person wide the *y* values can be interpreted as a probability.

Estimates for the mean value of *T*, over the different *I* values, as well as 95% confidence intervals can be derived from these distributions and are collected in Table III. Results are displayed for a value of ‘per 1000 students’ *i*.*e. N* = 1000.

**TABLE II:** Number and proportion of households in Wales containing at least one Higher Education student by household size. Source: Welsh Government analysis of the Annual Population Survey, 2019 [20].

| Number of people (including students) in a household | Number of households with at least one student | Proportion of all households containing at least one student (%) |
| --- | --- | --- |
| 1 | 8,300 | 8.3 |
| 2 | 22,000 | 22.1 |
| 3 | 27,600 | 27.6 |
| 4 | 27,700 | 27.7 |
| 5 | 10,100 | 10.1 |
| 6+ | 4,300 | 4.3 |
| Total | **99,900** | **100** |

**TABLE III:**
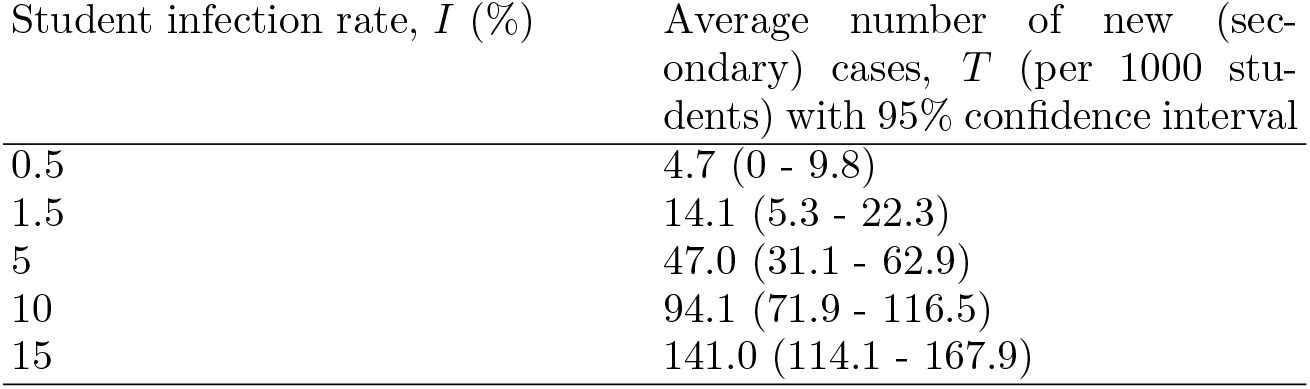
Mean average number of cases expected for varying levels of virus prevalence. Results based on 10^4^ repeated simulations of 1000 students.

| Student infection rate, $I$ (%) | Average number of new (secondary) cases, $T$ (per 1000 students) with 95% confidence interval |
| --- | --- |
| 0.5 | 4.7 (0 - 9.8) |
| 1.5 | 14.1 (5.3 - 22.3) |
| 5 | 47.0 (31.1 - 62.9) |
| 10 | 94.1 (71.9 - 116.5) |
| 15 | 141.0 (114.1 - 167.9) |

Considering the average number of secondary cases, we notice that a rough ‘rule of thumb’ for the expected number of secondary infections is each infected student will create just less than one further new case. For example, for a prevalence rate of 15% we would expect approximately 1000 × 0.15 = 150 secondary cases, which is within the error bounds for the simulated value of 141 secondary cases. Moreover, dividing each of the mean *T* values by the corresponding prevalences, *I*, and student population, *N*, provides a consistent probability of secondary infection (given an infected student) of *H* × *S* ≈ 0.94 (95% CI 0.69 – 1.19). Since *p* ≈ 1 this justifies our observation. Finally, considering equation (1) and using the mean vales from the literature we find that *T* ≈ 0.9*IN*, which again justifies the observation that each student infects just less than one other household member, on average.

## V. CONCLUSIONS

Using a Monte-Carlo based approach and current data from the Covid-19 literature, we have been able to predict that each infected student that is allowed to return home is expected to produce (on average) just less that one further secondary infection.

Critically, this work only accounts for transmissions and subsequent cases that might occur within a student’s household. We do not consider transmission to the wider home communities in which the students live. Further, we do not include the journey home, which may give rise to a larger number of cases, particularly if public transport is taken. Thus, our numbers are a lower bound on the likely impact of transmissions and new cases.

Until now, we have quoted the number of new cases per 1,000 students. However, using the average value of secondary infections to be 0.94 times the number of infected students, we can expand this out to the entire student population of Wales. Specifically, it is estimated that there are 99,900 households in Wales that contain at least one student in Higher Education. Although we do not readily have access to data on how many of these students live at home, versus those that live away and will return home for the holidays we can take the 99,900 as an upper bound. Thus, from our results we would expect approximately 4,670 (at a *I* = 5% infection level), or 1,409 (at a *I* = 1.5% infection level), new cases stemming from secondary infections from students returning home.

One also has to be mindful of household compositions and thus not just consider the total number of secondary cases, but who the students live with. From Welsh data [20] we know that 52.9% of all student households contain at least one other person with a diagnosed long-term illness. Such people are at greater risk of hospitalisation. Thus, for example from the additional 4,670 cases calculated above (at a 5% infection rate), 2,470 of these cases would be in those with a diagnosed long-term illness.

The accuracy of our data must be considered. Namely, the data we have used is either based on Cardiff University asymptomatic testing, Welsh Government analysis, or “FF100” (first few hundred) cases UK data. Thus, even if we can be confident in providing predictions for Welsh universities we urge the reader to rerun our simulations with their own data, which can be achieved using the Matlab code, or online app. The code and app are quick to run with a focus on accessibility so that a user can rapidly change the input probabilities to suit their data.

Universities cannot influence the makeup of a household. Equally, their influence on secondary infection rates in minimal. However, one potential suggestion is that universities could start their own media campaigns aimed at students’ households. For example they could send leaflets to a student’s registered home address that explains the risks that the household will face and steps that they can do to minimise their risk.

The only term within a university’s power to practically control is the prevalence of the virus in the student body, *I*. Critically, with the insight that every infected student that is allowed to leave is expected to infect at least one other person, it is imperative that plans are made to reduce *I* to as sustainably low as possible, as quickly as possible. Our work has already helped to influence government policy in relation to the firebreak (lockdown) in Wales during the period 23 October to 8 November 2020. Our model was used to assess the potential impact of individual, collective or institutional decisions for students to return from their halls of residence to their permanent home addresses. The analysis revealed that many of the students that would be affected live in multi-generational households and as a consequence this type of migration would expose older people living at the students’ home address to a higher level of risk of infection, ill-health and possible hospitalisation. Our analysis was considered by the Welsh Government’s Technical Advisory Cell and Higher Education Task and Finish Group dealing with Covid-19. On the basis of a consideration of the results of this work, Huw Morris (Director, Skills, Higher Education and Lifelong Learning, Welsh Government) confirmed that it was decided that there was evidence that encouraging students to return to their permanent home address from university residences would create greater risks than encouraging students to remain located close to their university of study. This decision and the reasoning behind it was communicated to colleagues in the UK Government, Scottish Government and Northern Ireland Executive and informed the wider development of policy in this area across the UK.

## VI. STRATEGIES

It is our hope that these findings inform the discussion on allowing students to travel back home safely over the Christmas period. Specifically, we now provide a range of suggested strategies and present their positive and negative aspects.

If infection rates are very low then secondary infection rates will also be low, thus, a university could attempt not to put any leaving restrictions in place. However, this does depend on predicting what infection rates may be in December, which is difficult. Equally, this strategy is not robust as it only requires a sudden outbreak in a hall of residence, for example, for the strategy to fail, resulting in many students having to self-isolate and thus not return home for Christmas. Or, more problematically, the students decide to return home in spite of the sudden outbreak and risk high numbers of secondary transmissions. Moreover even if prevalence rates are low, any asymptomatic student returning home is a threat to any vulnerable household occupants. Thus, if no student facing strategy is to be implemented we strongly advise key messaging to households, which might help raise awareness of risks of secondary household transmissions, especially for those living with vulnerable people.

Such a non-strategy on the part of a university could be backed up by home testing. Namely, on arrival back home, the student would have a test and a further followup test 5 days later. In the mean-time a student would need to self-isolate in their bedroom, at home, until the second test result is known. This would require shifting the responsibility onto the students, with expectations that they would not socialise during this time. This strategy would lower the transmission risks when at home, but it does not combat the risk during the initial travel, which is particularly troublesome for international students. This home-testing strategy also raises serious capacity implications for massive volumes of testing in a relative short space of time, although staggered departure dates could help reduce this pressure.

In contrast to having no strategy a university could bring in a strict lockdown. Namely, for the last two weeks of term all students are to self-isolate. The benefit of such a response is that the students and their household can be confident in their safe return. However, there are obvious serious student compliance and well-being issues. Indeed, such a strategy might cause students to leave their university accommodation early, before the lockdown’s implementation. The impact on a student’s health, both physical and mental has to be considered in such an extreme situation, specifically because many students will have already been in periods of self-isolation [3]. There will also be additional concerns for students on certain practice-based courses, *e*.*g*. medical and dental degrees, as they will miss important placements and clinical practice.

The requirements of a strict lockdown could be softened if on campus testing could be achieved. Namely, if students can be tested immediately before returning home then any students prompting a positive result would have to self-isolate for 10-14 days (hence return home a little later than planned). Alternatively, students prompting a negative result should ensure they only go onto campus for face to face teaching sessions and avoid all end of term socialising. Finally, they should have a second test the day before they go home. If negative then they go home, whilst if positive they are required to self-isolate. Under this campus-testing strategy less time is spent in self-isolation, perhaps prompting better compliance. Equally, the test results will reassure students and parents. However, compliance would still be an issue since we would require students to abstain from socialising during the run-up to Christmas. Equally, the ability to provide such a high volume of tests, as well as deliver results back in a timely manner, is a difficult logistic problem. However, as suggested with the home-testing strategy above, staggered departure dates could help reduce the pressure on the testing system.

Finally, if such campus-wide testing is not possible, but the prevalence is high enough that it is required then a university could focus its efforts on testing only those students who live with vulnerable people. This would dramatically reduce the amount of testing required, although it would cause new problems in estimating just how many tests would be needed. Equally, clear messaging to the students would be required to provide guidance on what counts as vulnerable.

The above suggestions are by no means a complete set of strategies. Neither are we suggesting that one plan of action will suit all universities due to their diversity of student body sizes and prevalence rates. Moreover, different universities maybe subject to different local governmental lockdown rules. The suggested strategies simply offer a broad set of guidelines that can be used and adapted, in conjunction with the data generated here, or by the reader generating their own data, to evidence a university’s specific chosen policy.

## Data Availability

All statistical data and processing codes are included in the article.

http://bit.ly/Secondary_infections_app

http://bit.ly/secondary_infections_repo

## Appendix A: Monte-Carlo Matlab code

The following code has been run and tested on Matlab R2019a. The code simulates `NS` (in our case 10^4^) stochastic samples of `N` students (in our case 1000). The code outputs the mean and 95% confidence intervals of the secondary infected population size.

The main inputs that can be modified by the user are: the probability distribution of household sizes, `Hhsizes`; the probabilities and standard deviations of secondary infections (depending on household size), `Mean_probs` and `Sd`, respectively; the number of simulated students, `N`; the number of runs to average over, `NS` and the prevalence rates, `I`.

Currently, the code includes data for household sizes ranging from 2 to 6, as this was what was available from the cited literature. However, should the user want to try alternative data for household sizes and second infection rates they must ensure that the vectors, `Hhsizes`, `Mean_probs` and `Sd` are all the same length.

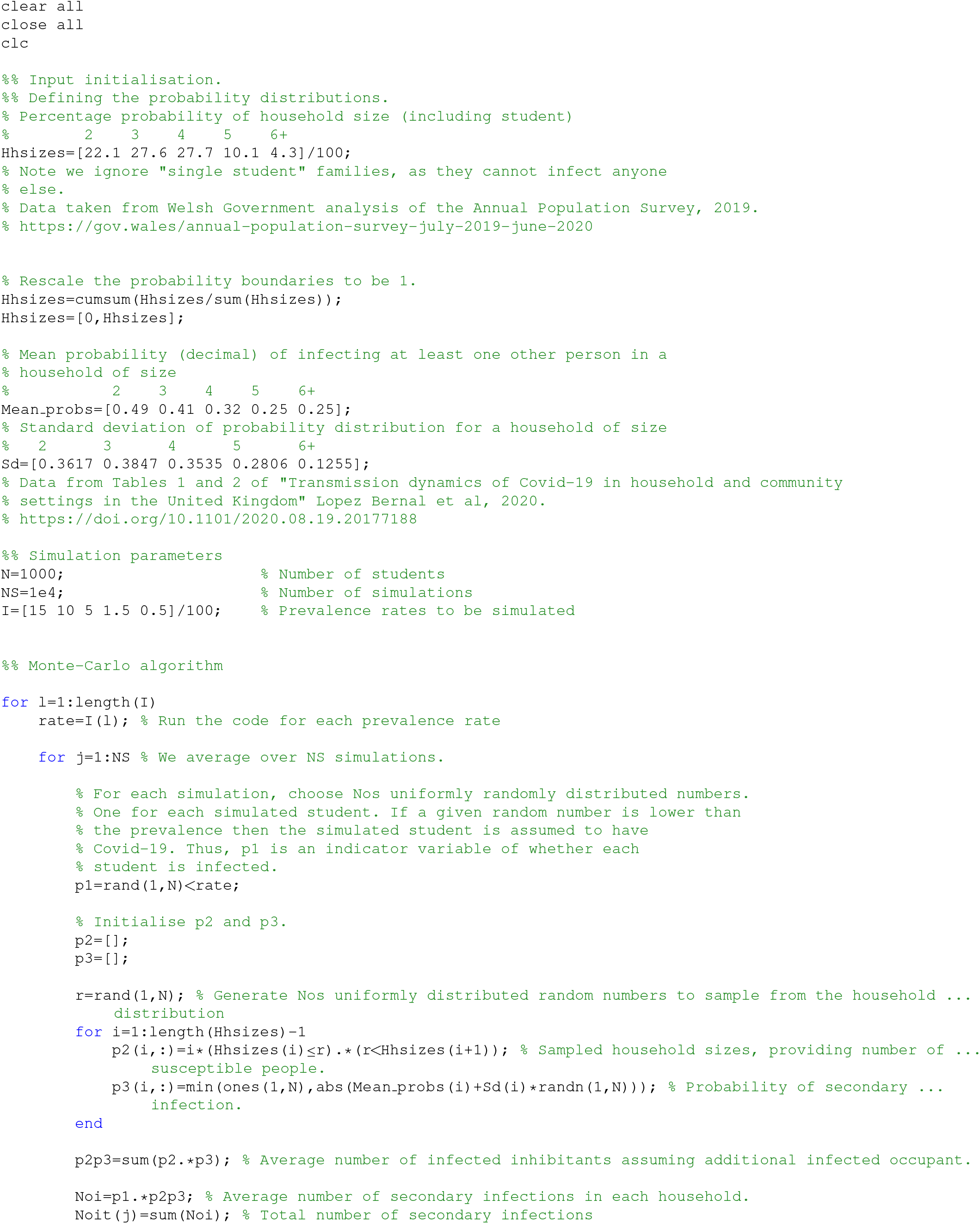
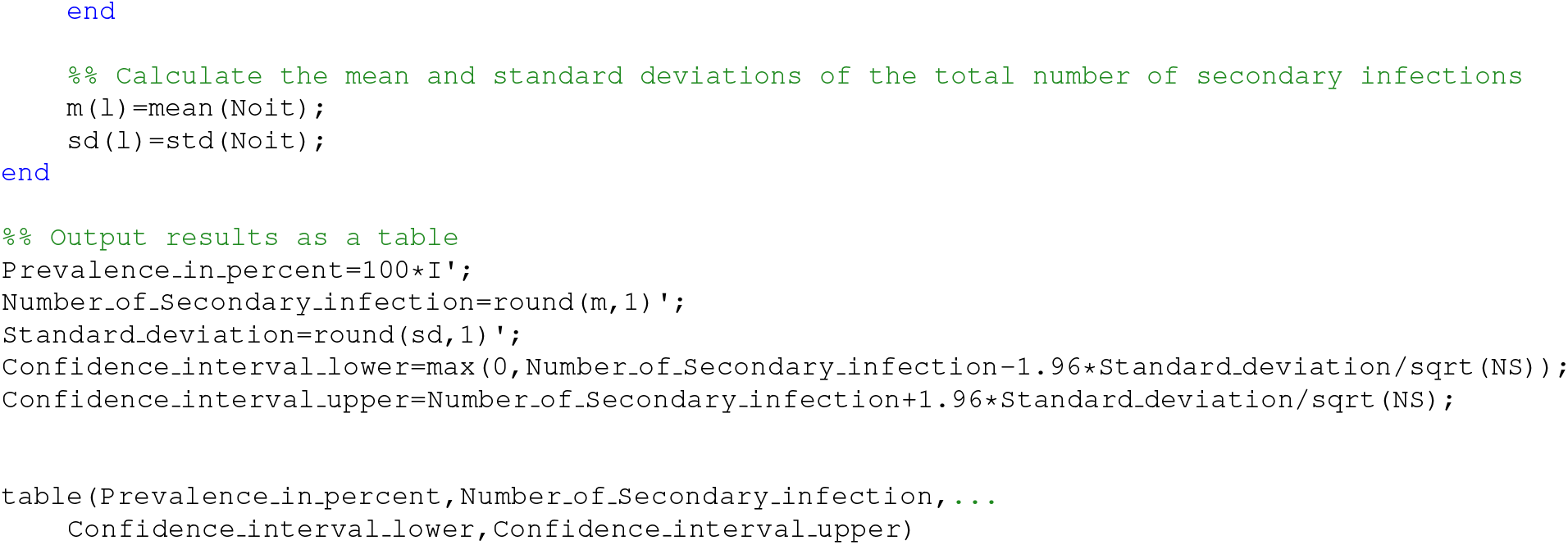

## References

[1] C. Wang, P. W. Horby, F. G. Hayden, and G. F. Gao. A novel coronavirus outbreak of global health concern. The Lancet, 395(10223):470–473, 2020.

[2] M. Chahrour, S. Assi, M. Bejjani, A. A. Nasrallah, H. Salhab, M. Fares, and H. H. Khachfe. A bibliometric analysis of Covid-19 research activity: A call for increased output. Cureus, 12(3), 2020.

[3] P. Sahu. Closure of universities due to Coronavirus Disease 2019 (COVID-19): impact on education and mental health of students and academic staff. Cureus, 12(4), 2020.

[4] C. Rapanta, L. Botturi, P. Goodyear, L Guardia, and M. Koole. Online university teaching during and after the Covid-19 crisis: Refocusing teacher presence and learning activity. Postdigi. Sci. Edu., pages 1–23, 2020.

[5] G. Yamey and R. P. Walensky. Covid-19: re-opening universities is high risk. Br. Med. J., 370, 2020.

[6] F. Perez-Reche and N. Strachan. Estimating the number of COVID-19 cases being introduced into UK Higher Education Institutions during Autumn 2020. medRxiv, 2020.

[7] R. A. Teran, I. Ghinai, S. Gretsch, T. Cable, S. R. Black, S. J. Green, O. Perez, G. E. Chlipala, M. Maienschein-Cline, and K. J. Kunstman. COVID-19 outbreak among a university’s men’s and women’s soccer teams—Chicago, Illinois, July–August 2020. MMWR Morb. Mortal Wkly Rep., 69:1591–1594, 2020.

[8] R. Laxminarayan, B. Wahl, S. R. Dudala, K. Gopal, C. Mohan B, S. Neelima, K. S. Jawahar Reddy, J. Radhakrishnan, and J. A. Lewnard. Epidemiology and transmission dynamics of COVID-19 in two Indian states. Science, 370(6517): 691–697, 2020.

[9] https://www.euro.who.int/en/health-topics/health-emergencies/coronavirus-covid-19/statements/statement-older-people-are-at-highest-risk-from-covid-19-but-all-must-act-to-prevent-community-spread. WHO Statement – Older people are at highest risk from COVID-19, but all must act to prevent community spread, Accessed 4th November, 2020.

[10] Britton, F. Ball, and P. Trapman. A mathematical model reveals the influence of population heterogeneity on herd immunity to SARS-CoV-2. Science, 369(6505):846–849, 2020.

[11] J. Ripperger, J.L. Uhrlaub, M. Watanabe,R. Wong, Y. Castaneda, H. A. Pizzato, M. R. Thompson, C. Bradshaw, C. C. Weinkauf, C. Bime, H. L. Erickson, K. Knox, B. Bixby, S. Parthasarathy, S. Chaudhary, B. Natt, E. Cristan, T. El Aini, F. Rischard, J. Campion, M. Chopra, M. Insel, A. Sam, J. L. Knepler, A. P. Capaldi, C. M. Spier, M. D. Dake, Edwards, M. E. Kaplan, S. J. Scott, C. Hypes, J. Mosier, D. T. Harris, B. J. LaFleur, R. Sprissler, J. Nikolich-Žugich, and D. Bhattacharya. Orthogonal SARS-CoV-2 Serological Assays Enable Surveillance of Low-Prevalence Communities and Reveal Durable Humoral Immunity. Immunity, 2020.

[12] W. Tan, Y. Lu, J. Zhang, J. Wang, Y. Dan, Z. Tan, X. He, C. Qian, Q. Sun, and Q. Hu. Viral kinetics and antibody responses in patients with COVID-19. medRxiv, 2020.

[13] K. A. Callow, H. F. Parry, M. Sergeant, and D. A. J. Tyrrell. The time course of the immune response to experimental coronavirus infection of man. Epidem. Infect., 105(2):435–446, 1990.

[14] J. Seow, C. Graham, B. Merrick, S. Acors, S. Pickering, O. Steel, K. J. A .and Hemmings, A. O’Byrne, N. Kouphou, and R. P. Galao. Longitudinal observation and decline of neutralizing antibody responses in the three months following SARS-CoV-2 infection in humans. Nature Microbiol., pages 1–10, 2020.

[15] M. M. Husky, V. Kovess-Masfety, and J. D Swendsen. Stress and anxiety among university students in France during Covid-19 mandatory confinement. Compre. Psych., 102:152191, 2020.

[16] H. Christensen, K. Turner, A. Trickey, R. D. Booton, G. Hemani, E. Nixon, C. Relton, L. Danon, M. Hickman, and E. B. Pollock. COVID-19 transmission in a university setting: a rapid review of modelling studies. medRxiv, 2020.

[17] https://www.ons.gov.uk/peoplepopulationandcommunity/healthandsocialcare/conditionsanddiseases/articles/coronaviruscovid19infectionsinthecommunityinengland/october2020. Coronavirus (COVID-19) Infection Survey: characteristics of people testing positive for COVID-19 in England: October 2020, accessed 4th November.

[18] https://www.cardiff.ac.uk/coronavirus/covid-19-casenumbers. COVID-19 case numbers in Cardiff University, Accessed 4th November.

[19] J. Lopez Bernal, N. Panagiotopoulos, C. Byers, T. Garcia Vilaplana, N. L. Boddington, X. Zhang, A. Charlett, S. Elgohari, L. Coughlan, R. Whillock, S. Logan, H. Bolt, M. Sinnathamby, L. Letley, P. MacDonald, R. Vivancos, O. Edeghere, C. Anderson, K. Paranthaman, S. Cottrell, J. McMenamin, M. Zambon, G. Dabrera, M. Ramsay, and V. Saliba. Transmission dynamics of COVID-19 in household and community settings in the United Kingdom. medRxiv, 2020.

[20] https://gov.wales/annual-population-survey-july-2019-june 2020. Annual Population Survey: July 2019 to June 2020, accessed 4th November.

[21] D. P. Landau and K. Binder. A Guide to Monte Carlo Simulations in Statistical Physics. Cambridge University Press, 2014.

[22] J. Wu, Y. Huang, C. Tu, C. Bi, Z. Chen, L. Luo, M. Huang, M. Chen, C. Tan, Z. Wang, K. Wang, Y. Liang, J. Huang, X. Zheng, and J. Liu. Household Transmission of SARS-CoV-2, Zhuhai, China, 2020. Clin. Infect. Dis., 2020.

[23] E. S. Rosenberg, E. M. Dufort, D. S. Blog, E. W. Hall, D. Hoefer, B. P. Backenson, A. T. Muse, J. N. Kirkwood, K. St. George, D. R. Holtgrave, B. J. Hutton, and H. A. Zucker. COVID-19 Testing, Epidemic Features, Hospital Outcomes, and Household Prevalence, New York State—March 2020. Clin. Infect. Dis., 71(8):1953–1959, 2020.

[24] N. M. Lewis, V. T. Chu, D. Ye, E. E. Conners, R. Gharpure, R. L. Laws, H. E. Reses, B. D. Freeman, M. Fajans, E. M. Rabold, P. Dawson, S. Buono, S. Yin, D. Owusu, A. Wadhwa, M. Pomeroy, A. Yousaf, E. Pevzner, H. Njuguna, K. A. Battey, C. H. Tran, V. L. Fields, P. Salvatore, M. O’Hegarty, J. Vuong, R. Chancey, C. Gregory, M. Banks, J. R. Rispens, E. Dietrich, P. Marcenac, A. M. Matanock, L. Duca, A. Binder, G. Fox, S. Lester, L. Mills, S. I. Gerber, J. Watson, A. Schumacher, L. Pawloski, N. J. Thornburg, A. J. Hall, T. Kiphibane, S. Willardson, K. Christensen, L. Page, S. Bhattacharyya, T. Dasu, A. Christiansen, I. W. Pray, R. P. Westergaard, A. C. Dunn, J. E. Tate, S. A. Nabity, and H. L. Kirking. Household Transmission of SARS-CoV-2 in the United States. Clin. Infect. Dis., 2020.

